# Loss-of-function Variants in *CPT1C*: No Support for a Causal Role in Hereditary Spastic Paraplegia

**DOI:** 10.1101/2025.09.18.25335173

**Authors:** Rui Zhu, Lang Liu, Mehrdad A Estiar, Farnaz Asayesh, Jamil Ahmad, Meron Teferra, Grace Yoon, Mark Tarnopolsky, Kym M Boycott, Nicolas Dupre, Patrick A Dion, Oksana Suchowersky, Albena Jordanova, Yi-Chung Lee, Giovanni Stevanin, Stephan Zuchner, Guy A Rouleau, Ziv Gan-Or

## Abstract

**Background:** Hereditary spastic paraplegias (HSPs) are neurodegenerative disorders characterized by lower limb spasticity. Pathogenic variants in *CPT1C* have been implicated in HSP.

**Objective:** To assess if *CPT1C* loss-of-function (LOF) variants are causally associated with HSP.

**Methods:** We analyzed whole-genome sequencing (WGS) data from UK Biobank (UKBB), whole-exome sequencing (WES) data from a Canadian cohort of HSP (Can-HSP), and genetic data from the GENESIS cohort—a large international cohort of patients with rare hereditary diseases, including HSP.

**Results:** Among >170 *CPT1C* LOF carriers in the UKBB (n=150,119), none exhibited HSP phenotypes. Among 585 HSP patients from Can-HSP, we did not find patients with *CPT1C* LOF variants. In the GENESIS cohort (n=21,217), three individuals carrying *CPT1C* LOF variants were also diagnosed with HSP; however, all three also carry pathogenic variants in established HSP-associated genes.

**Conclusion:** Our study does not support a causal role for *CPT1C* LOF variants in HSP.

## INTRODUCTION

Hereditary spastic paraplegias (HSPs) are a group of heterogeneous neurodegenerative disorders characterized primarily by progressive lower limb spasticity and weakness1. The prevalence of HSP ranges from 3-10 cases per 100,000 individuals (0.003% to 0.01%)^2, 3^. Over 80 HSP-associated genes have been identified to date, including all major inheritance patterns: autosomal dominant, autosomal recessive, X-linked, and mitochondrial forms4.

Autosomal dominant HSP is the most common inheritance pattern of HSP, accounting for approximately 75%-80% of all cases. Over the last decade, five studies have implicated variants in *CPT1C* as the cause of SPG73, a rare autosomal dominant subtype of HSP, in 14 affected individuals from seven unrelated families5-9. CPT1C, Carnitine palmitoyltransferase 1C, regulates lipid metabolism and energy homeostasis and is mainly localized in the endoplasmic reticulum of neurons10. The first identified *CPT1C* variant in HSP was a missense mutation that affected lipid metabolism in human motor neurons5 while subsequent reports described loss-of-function (LOF) variants6-9. Based on these studies, it was suggested that LOF of *CPT1C* may lead to HSP. However, the evidence supporting a direct causal relationship remains limited.

In this study, we thoroughly evaluate the association of *CPT1C* LOF variants with HSP using data from the UK Biobank (UKBB), a Canadian HSP cohort (Can-HSP)11, and the GENESIS database—an international rare disease database enriched for patients with hereditary neurogenetic disorders, including HSP12.

## METHODS

### Population and Genetic Datasets

To assess whether *CPT1C* LOF variants are causally associated with HSP, we analyzed three datasets: UKBB, Can-HSP, and GENESIS: *UKBB:* We obtained genetic data from the UKBB GraphTyper whole-genome sequencing (WGS) dataset (n=150,119). Sequence alignment was performed using the GRCh38/hg38 human reference genome13. *Can-HSP*: A total of 585 HSP patients from 372 families were recruited across Canada. Of these, 400 HSP patients underwent whole-exome sequencing (WES). WES reads were aligned to the GRCh37/hg19 human reference genome11. *GENESIS*: We accessed the GENESIS dataset (n=21,217), which includes datasets from individuals with rare monogenic diseases, such as HSP12.

### Variant Filtering and Prioritization

The UKBB GraphTyper WGS data was pre-filtered in UKBB using a logistic regression model that retains only variants with AAscore greater than 0.5. The AAscore is a confidence score representing the predicted probability that a variant is a true positive13. All the *CPT1C* LOF variants we extracted from UKBB had AAscores ≥ 0.8997, suggesting a very high confidence in variant calls. All variants in WES data from the Can-HSP underwent standard quality control as previously reported11, and only variants that passed all the filtering steps were used as input for annotation with Annotate Variation (ANNOVAR). In the GENESIS database, we prioritized high-confidence, rare, and likely pathogenic variants. We included variants predicted to be LOF or damaging by Variant Effect Predictor (VEP): *transcript ablation, splice acceptor, splice donor, stop-gained, frameshift, start-loss*, and *stop-loss*. Given the low prevalence of HSP, and to restrict our analysis to rare alleles, we applied a maximum allele frequency of Genome Aggregation Database (gnomAD) v2 Allele Frequency (AF) ≤ 1 × 10^−5^ . To enrich for likely pathogenic variants, we filtered variants with a Combined Annotation Dependent Depletion (CADD) score of > 20. We also filtered variants to ensure high sequencing quality, applying the following criteria: read depth ≥ 15 for short reads (SR), ≥ 10 for long reads (LR); genotype quality of ≥ 75 for SR, and ≥ 30 for LR, and a variant quality score of ≥ 250.

### Variant Annotation and Analysis

To assess the potential causal relationship between *CPT1C* LOF mutations and HSP, we analyzed the phenotypic data from individuals in the UKBB carrying such variants. We performed variant annotation using both ANNOVAR14 and VEP15 on the GRCh38/hg38 reference genome to identify LOF variants in *CPT1C*.

WES data from the Can-HSP cohort were annotated using ANNOVAR and aligned to the GRCh37/hg19 reference genome11. In the UKBB datasets and the GENESIS database, we first identified individuals carrying *CPT1C* LOF variants and then evaluated their phenotypes and diagnoses. In the Can-HSP cohort, which consists entirely of HSP patients, we assessed whether these patients carried *CPT1C* LOF variants.

Since the GENESIS cohort was the only dataset in which we identified individuals with both HSP and *CPT1C* LOF variants, we applied the American College of Medical Genetics and Genomics (ACMG) guidelines16 to these variants, along with those reported in previously published studies6-9.

## DATA SHARING

Information relevant to the analyses is available in the supplementary file of this article.

## RESULTS

### Individuals from UKBB with *CPT1C* LOF mutations do not show HSP

Using ANNOVAR, we identified 29 *CPT1C* LOF variants in 171 individuals. We obtained their associated phenotypic data from UKBB. Analysis revealed 471 unique phenotypes among carriers, none associated with HSP (Supplemental Table 1). The five most common phenotypes among these individuals were cataract (n=29), hernia (n=19), diverticular disease (n=15), abdominal pain (n=13), and gonarthrosis (n=11, Supplemental Table 2).

**Table 1.** Genetic and Inheritance Patterns of HSP Families from GENESIS with *CPT1C* Variants.

| HSP Diagnosis | Known HSP gene variant(s) (HGVS formatting) | *CPT1C* variant (HGVS formatting) | Inheritance Pattern |
| --- | --- | --- | --- |
| Complex HSP | NM_015346.4:c.3523+1del and NM_015346.4:c.1272-14A>G | NM_001199753.2:c.1018C>T | Recessive (ZF2VE26) |
| Autosomal recessive<br>SPG15 | NM_015346.4:c.3523+1del and NM_015346.4:c.1272-14A>G | NM_001199753.2:c.1018C>T | Recessive (ZF2VE26) |
| Pure HSP | SPAST exon 17 deletion (MLPA-confirmed) | NM_001199753.2:c.345G>A | Dominant (SPAST) |

**Table 2.**
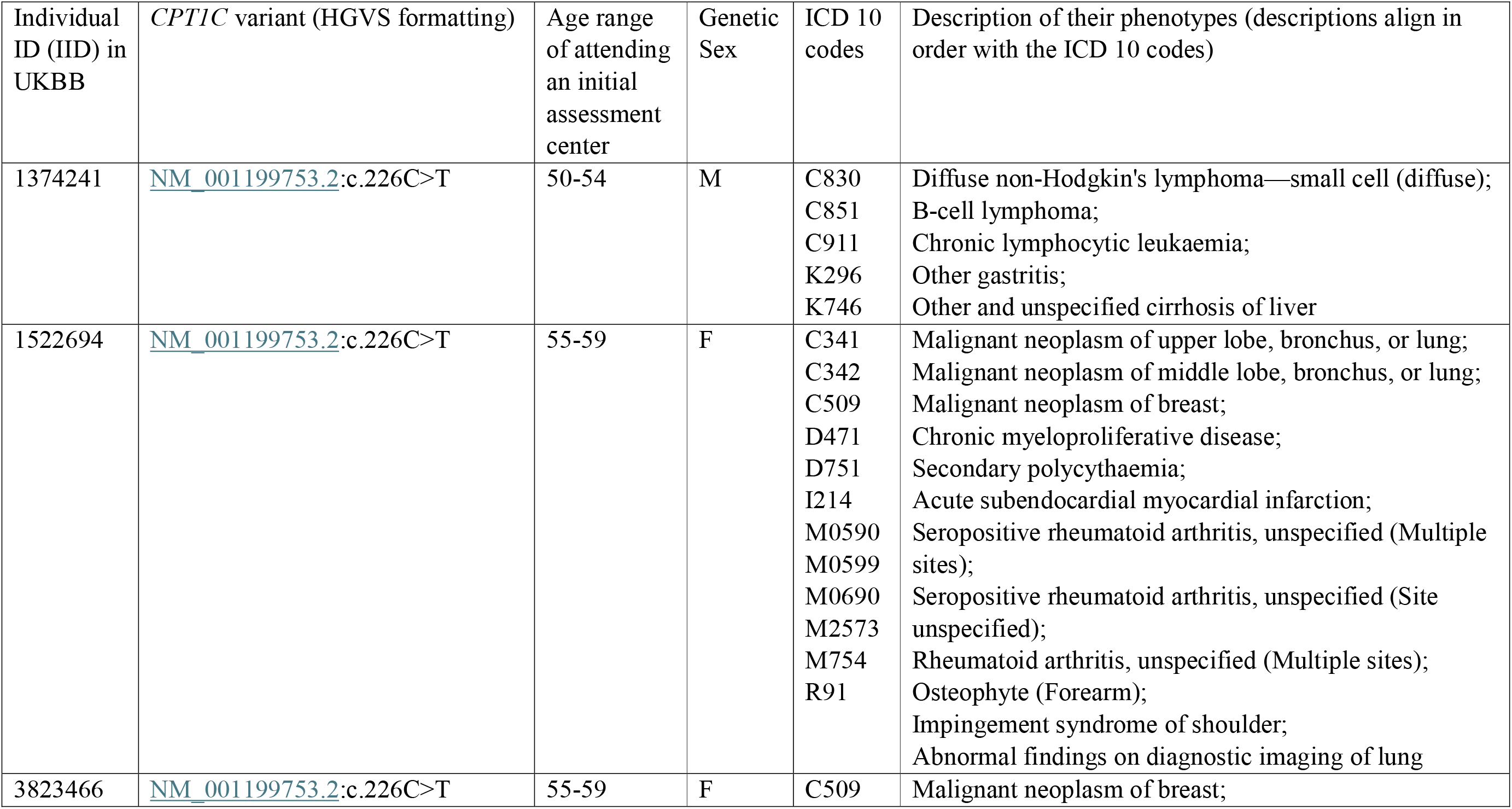
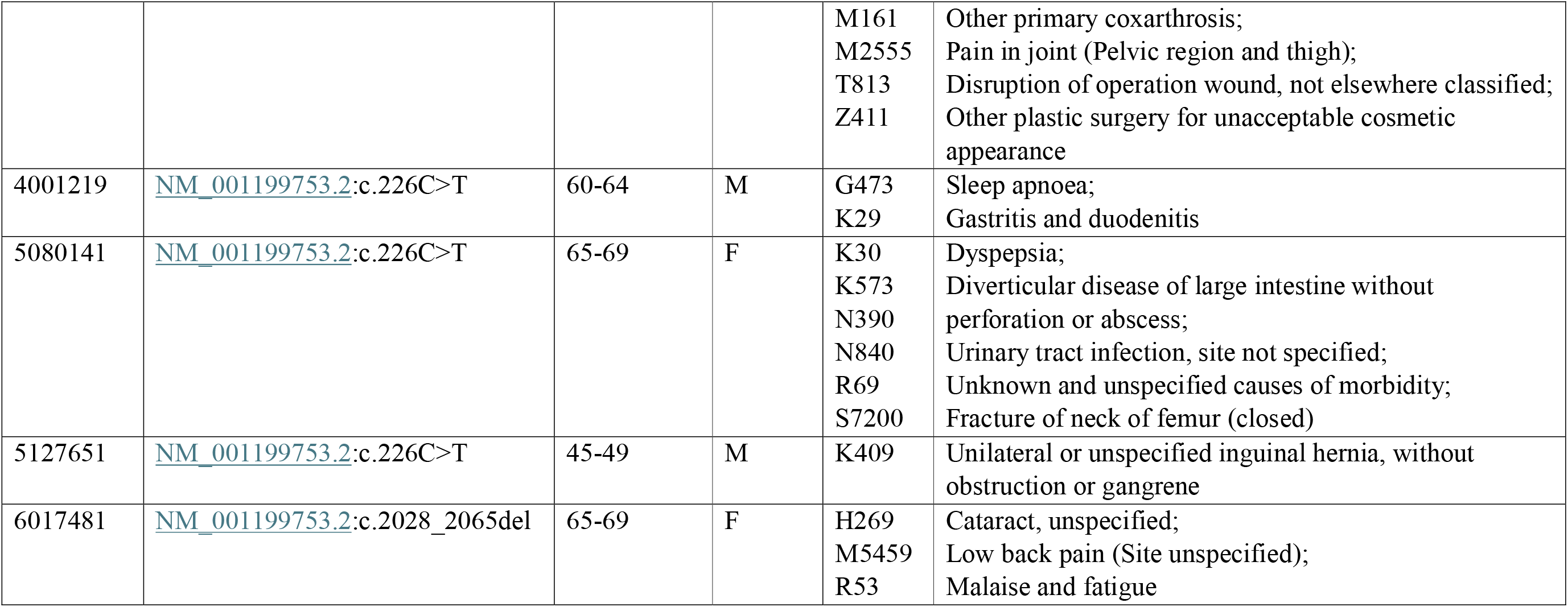
Phenotypes displayed by UKBB individuals with *CPT1C* LOF variants (<u>NM_001199753.2</u>:c.226C>T; <u>NM_001199753.2</u>:c.2028_2065del)

Using VEP, we identified 55 *CPT1C* LOF variants, found in 225 individuals. Analysis of these carriers revealed a total of 597 distinct phenotypes, none related to HSP (Supplemental Table 3). The five most common phenotypes observed among these individuals were similar to those above, with cataract being the most common (n=35), followed by hernia (n=28), abdominal pain (n=26), diverticular disease (n=20), and gonarthrosis (n=16, Supplemental Table 4).

### Analysis of *CPT1C* in Can-HSP and GENESIS

We did not identify any HSP patients from the Can-HSP cohort with *CPT1C* LOF variants. Among 21 individuals in the GENESIS cohort carrying *CPT1C* LOF variants, three were diagnosed with HSP. We further followed up on the precise genetic diagnoses for HSP in these individuals.

One individual comes from a family with two healthy parents and two affected children, suggesting a recessive inheritance pattern. Compound heterozygous variants in the *ZFYVE26* gene (SPG15)17, were found in the affected family members: <u>NM_015346.4</u>:c.3523+1del, abolishing the wild type splice site at the exon19-intron19 junction18, and <u>NM_015346.4</u>:c.1272-14A>G, creating a novel splice site with higher splice site scores compared to the wild type one in the intron8-exon9 junction18. Another unrelated individual was also diagnosed with autosomal recessive SPG1517. The third individual was diagnosed with HSP due to a pathogenic deletion of exon 17 in *SPAST* (Table 1).

## DISCUSSION

Overall, our findings suggest that LOF variants in *CPT1C* are not associated with HSP. In UKBB, we identified 171 and 225 individuals carrying *CPT1C* LOF variants among 150,119 whole-genome sequenced individuals using ANNOVAR and VEP, respectively. These prevalences, of 0.11% and 0.15%, are 10-50 times higher than the known prevalence of HSP in Europeans2, suggesting that is unlikely that *CPT1C* LOF variants cause HSP. Furthermore, none of these individuals were diagnosed with HSP or presented with HSP-related phenotypes.

The three individuals in the GENESIS cohort also had alternative HSP genetic diagnoses: two carried variants in *ZFYVE26*, and the other had a pathogenic deletion of exon 17 in *SPAST*. As all three individuals with *CPT1C* LOF variants also harbored pathogenic mutations in genes already implicated in well-known HSP subtypes^17, 19, 20^, their clinical presentations are more plausibly explained by these established genetic causes.

We applied the ACMG criteria to assess the pathogenicity of CPT1C *LOF* variants16. The unique *CPT1C* LOF variants identified using ANNOVAR and VEP in the UKBB cohort were observed in individuals with a mean age of 57.5 and 56.7 years, respectively, with the youngest being 40 years old at the initial assessment. In contrast, prior studies reported HSP associated with *CPT1C* LOF variants manifesting in infancy or childhood6-8 (Supplemental Table 5). Thus, the variants met ACMG criterion (Benign Strong 2) BS2—observed in healthy adults in the context of a disorder that is expected to be fully penetrant at an early age16, supporting strong evidence of benign impact of these *CPT1C* LOF variants on HSP. Additionally, the three carriers of *CPT1C* LOF variants in the GENESIS database had confirmed alternate genetic diagnoses of HSP, thus satisfying ACMG criterion Benign Supporting 5 (BP5)16, suggesting that *CPT1C* variants in these cases are likely incidental findings.

In previously reported cases 6-9 (Supplemental Table 5), segregation data are insufficient. For instance, the *CPT1C* variant (<u>NM_001199753.2</u>:c.226C>T)6 was inherited from an asymptomatic mother who exhibited no HSP-related symptoms prior to her daughter’s diagnosis. Similarly, in two other cases 7, both patients had LOF mutations in the *CPT1C* gene (<u>NM_001199753.2</u>:c.226C>T; <u>NM_001199753.2</u>:c.524_527dup), inherited from asymptomatic parents. Both index patients demonstrated HSP-related symptoms in infancy, yet the healthy adult carriers again meet ACMG criterion BS216, arguing against the pathogenicity of *CPT1C* LOF variants in HSP. Another case 8 involved a maternally inherited *CPT1C* variant (<u>NM_001199753.2</u>:c.2020-1G>C), yet the mother exhibited only weak, atypical symptoms. In a recent case9, only the index patient and her mother were genotyped and found with the *CPT1C* variant (<u>NM_001199753.2</u>:c.310del), while the other affected family members were not genotyped, leaving open the possibility of non-segregation. These cases do not provide compelling evidence for *CPT1C* LOF variants being pathogenic and causal for autosomal dominant HSP. We found six individuals with the *CPT1C* variant reported in two studies ^6, 7^ <u>NM_001199753.2</u>:c.226C>T) in UKBB. None of the six individuals displays HSP phenotypes (Table 2), providing strong population-level evidence against the pathogenicity of this *CPT1C* LOF variant in HSP.

Our study has several limitations. The phenotype data in UKBB may lack the resolution to detect early-stage neurological symptoms, and no imaging phenotyping or electrophysiological studies were accessible in the carriers. Indeed, as UKBB participants are middle-aged to older adults, some signs of mild or pure spastic paraplegia may have been missed. While our findings strongly argue against a monogenic causal role of *CPT1C* LOF variants in HSP, we cannot exclude the possibility that these variants may act as modifiers or contribute to disease in the context of additional genetic or environmental factors. An incomplete penetrance or variable expressivity of LOF variants is also possible.

In conclusion, while *CPT1C* LOF variants have been reported in individuals with HSP, our analysis across population-scale data (UKBB), HSP-specific cohort (Can-HSP), and disease-specific cohorts (GENESIS) reveals no consistent evidence for a causal role of *CPT1C* LOF variants in HSP. The absence of phenotype in adult carriers, the presence of alternative HSP diagnoses, and the weak segregation undermine the proposed pathogenicity of *CPT1C* LOF variants in SPG73. These findings support reclassifying *CPT1C* LOF variants as likely benign in the context of HSP in the absence of functional elements towards their implication in SPG73.

## Data Availability

All data produced in the present study are available upon reasonable request to the authors

## ACKNOWLEDGMENT

We thank the patients and their families for participating in this study. This study was funded by CIHR Emerging Team Grant, in collaboration with the Canadian Organization for Rare Disorders (CORD), grant number RN127580-260005, and by a CIHR Foundation grant granted to GAR. This research was undertaken thanks in part to funding from the Canada First Research Excellence Fund, awarded to McGill University for the Healthy Brains for Healthy Lives initiative granted to MAE.

## ETHICS STATEMENT

For Can-HSP, all participants have signed an informed consent form prior to enrollment, and the McGill University Health Center (MUHC) Research Ethics Board (REB) has approved the study protocols, REB number IRB00010120. IRB of GENESIS does not apply. No human subject data was involved. Consent of participants happened at sites that submit data.

## AUTHORS’ ROLES

1. Research project: A. Conception, B. Organization, C. Execution;
2. Statistical Analysis: A. Design, B. Execution, C. Review and Critique;
3. Manuscript Preparation: A. Writing of the first draft, B. Review and Critique R.Z.: 1A, 1B, 1C, 2A, 2B, 3A.

L.L.: 1C, 2C, 3B

M.E.: 1C, 2C, 3B

F.A.: 1C

J.A.: 1C

M.T.: 1C

G.Y.: 2C, 3B

M.T.: 2C, 3B

K.B.: 2C, 3B

N.D.: 2C, 3B

P.D.: 2C, 3B

O.S.: 2C, 3B

A.J.: 2C, 3B

Y.L.: 2C, 3B

G.S.: 2C, 3B

S.Z.: 2C, 3B

G.R.: 2C, 3B

Z.G.-O.: 1A, 1B, 2C, 3B

## FINANCIAL DISCOLSURES OF ALL AUTHORS OF THE PRECEDING 12 MONTHS

ZGO has received consulting fees from Lysosomal Therapeutics Inc., Idorsia, Prevail Therapeutics, Denali, Ono Therapeutics, Neuron23, Handl Therapeutics, UBC, Bial Biotech Inc., Bial, Deerfield, Guidepoint, Lighthouse and VanquaBio. MK is the CEO of Exerkine Corporation. He has received financial compensation from Amicus Therapeutics for work on an Ad board in 2025. ND has received funding from Centre de recherche du CHU de Québec – Université Laval, Association de la neurofibromatose du Québec (ANFQ), Plateforme de recherche clinique du CHU de Québec – Université Laval, ALZHEIMER’S DRUG DISCOVERY FOUNDATION, Fonds de recherche du Québec - Santé (FRQS), and The Royal Institution for the Advancement of Clinical research. OS has received grants from WaveLifeSciences, Roche, Alnylam, related to Hungtington Disease. GS received grants from The Association Strumpell Lorrain HSP-France, from the Agence Nationale de la Recherche (NEURO-NGS3) and from the Programme d’Investissements d’Avenir Idex Université de Bordeaux. GS’s work was financed by the Association Connaitre les syndromes cérébelleux, and by Campus France (PHC PESSOA). None of these companies were involved in any parts of preparing, drafting and publishing this study. Other authors have nothing to disclose.

## SUPPLEMENTAL TABLE LEGENGS

**Supplemental Table 1**. The phenotypic data of the 171 individuals carrying *CPT1C* LOF variants in UKBB, annotated by ANNOVAR.

**Supplemental Table 2**. Top Five Phenotypes Observed in UKBB Individuals Carrying *CPT1C* LOF Variants Identified via ANNOVAR

**Supplemental Table 3**. The phenotypic data of the 225 individuals carrying *CPT1C* LOF variants in UKBB, annotated by VEP.

**Supplemental Table 4**. Top Five Phenotypes Observed in UKBB Individuals Carrying *CPT1C* LOF Variants Identified via VEP

**Supplemental Table 5**. Summary of the phenotypes of HSP patients carrying *CPT1C* LOF variants from previous studies.

